# Factors associated with maternal tetanus vaccination in Myanmar: An analysis of Demographic and Health Survey data

**DOI:** 10.1101/2021.03.08.21253169

**Authors:** Zaw Myo Tun, Zau Ring, Clarence C Tam

**Affiliations:** Saw Swee Hock School of Public Health, National University of Singapore, 12 Science Drive 2, #10-01, Singapore 117549; Vector Borne Diseases Control Program, Kachin State Public Health Department, Ministry of Health and Sports, Myanmar Kachin State Public Health Department Office, Yone Gyi Street, Ayar Quarter, Myitkyina, Myanmar; London School of Hygiene & Tropical Medicine, Keppel Street, London WC1E 7HT

**Keywords:** Tetanus toxoid vaccination, Myanmar, Universal health coverage, Epidemiology, Demographic Health Survey

## Abstract

**Background:** Tetanus toxoid vaccination is a life-saving maternal and child health intervention. Understanding gaps in maternal vaccination coverage is key to informing progress towards universal health coverage. We assessed maternal tetanus vaccination coverage in Myanmar and investigated factors associated with being unvaccinated.

**Method:** We analysed 2015-16 Demographic and Health Survey data including women aged 15-49 years with at least one childbirth in the last five years. The outcome was self-reported receipt of tetanus vaccine at least once during the last pregnancy. We used logistic regression models to assess factors associated with being unvaccinated.

**Results:** Overall maternal tetanus vaccination coverage was 85.7%. Sub-national coverage was lowest in the predominantly ethnic minority states of Shan, Kayin, and Chin at 69.6%, 77.4%, and 79.9%, respectively. Factors associated with a lack of vaccination were: not receiving antenatal care (odds ratio (OR): 18.99, 95% confidence interval (CI): 14.21, 25.39); receiving antenatal care at home (OR: 2.05, 95% CI: 1.46, 2.88), private and non-governmental organization clinics (OR: 2.88, 95% CI: 1.81, 4.58, compared to public facilities); and not wanting to go to a health facility alone (OR: 1.53, 95% CI: 1.14, 2.06). Higher educational attainment was associated with lower odds of being unvaccinated (OR: 0.48, 95% CI: 0.32, 0.70 for secondary relative to no education).

**Interpretation:** We identified regional, structural, and individual differences in maternal tetanus vaccination coverage. Achieving universal coverage of maternal tetanus vaccination will largely depend on the ability to provide accessible antenatal care to most women who do not currently receive it.

## Introduction

Maternal and neonatal tetanus (MNT) is a devastating disease with a high case fatality, especially in low-income settings [1]. The World Health Organization (WHO) estimated 25,000 new-borns died from neonatal tetanus in 2018 [1]. MNT remains a significant public health problem in 12 countries, most of them are in Asia and Africa [1]. MNT death is preventable by hygienic delivery, cord care practices, and by immunizing children and pregnant women with inexpensive and efficacious tetanus toxoid vaccine [1]. In the last 30 years, vaccinating pregnant women in their second or third trimester has reduced tetanus-related neonatal mortality by 94% [2]. WHO recommends that “all women giving birth and their newborn babies should be protected against tetanus” and that countries should ensure that a national policy and strategies to achieve high vaccination coverage among pregnant women are available and are correctly implemented [3].

In Myanmar, a population of 51.5 million [4], tetanus vaccination for pregnant women has been part of the routine Expanded Program on Immunization since 1978 [5]. Although MNT was eliminated (defined as <1 MNT per 1000 live births in every district) in Myanmar in 2010 [6], the MNT burden and gaps in MNT preventive services remains: 231 neonatal tetanus cases were reported from 2011 to 2018, and 64% of mothers of neonatal tetanus cases in 2018 were unimmunized [7]; only 72% of women giving birth in the previous five years are protected against neonatal tetanus [5].

Factors associated with maternal tetanus vaccination are not well understood in Myanmar. The Myanmar Ministry of Health and Sports launched the five-year National Health Plan in December 2016 to achieve universal health coverage by 2030, aiming to ensure access to a basic essential package of health services by the entire population [8]. Maternal tetanus vaccination is listed as a component of the essential package [9]. Information on factors associated with maternal tetanus under-vaccination would be useful to inform the implementation strategy. We estimated maternal tetanus vaccination coverage at both national and subnational levels using 2015-16 Myanmar Demographic and Health Survey (MDHS) data. We also investigated factors associated with being unvaccinated among women reporting pregnancy in the previous five years.

## Methods

We used maternal health data from the 2015-16 MDHS which was implemented by the Ministry of Health and Sports with support from the Inner City Fund (ICF) (Rockville, Maryland, USA). The DHS collected data on demographic and health indicators of women and their household members using a nationally representative sample. The survey methodology and data collection procedures have been published in the main report [5]. Briefly, the survey design employed a two-stage sampling (441 clusters, 30 households per cluster) stratified by urban and rural areas in 15 subnational administrative divisions (seven states, seven regions, and Nay Pyi Taw Union Territory). In this analysis, Nay Pyi Taw Union Territory was categorized as a region. The states, representing the mountainous areas, are the areas of residence for seven ethnic minority groups while the majority Burmese ethnicity primarily live in the eight regions which represent the plain areas. The states are generally less developed than the regions [4]. In this cross-sectional analysis, we included women aged 15-49 years who reported giving birth in the last five years.

### Outcome variable

Outcome was being unvaccinated with tetanus toxoid containing vaccine (TTCV) self-reported by mothers. Maternal tetanus vaccination was defined as newborns protected against neonatal tetanus (See Appendix 1 for the full definition) according to the Myanmar National Guidelines for Antenatal Care [10]. The following variables were used to construct the outcome variable: (1) number of tetanus injection before birth (during the last pregnancy); (2) number of tetanus injections before the last pregnancy; (3) number of years ago received last tetanus injection before the last pregnancy. We estimated the percentage of unvaccinated mothers with corresponding 95% confidence interval (CI), at national and subnational levels, accounting for the DHS sampling weights. We also compared vaccine coverage with the percentage of mothers who received at least one dose of TTCV during their last pregnancy in each state and region. Their differences indicate mothers who missed a second dose of TTCV as the vast majority of vaccinated mothers received two TTCV injections (See results section below for more details).

### Explanatory variables

We selected 13 variables and grouped them into three domains: (1) Socio-demographic: residence in a state or a region, urban or rural residence, age (categorized as 15-24, 25-29, 30-34, 35-49), birth order of the last pregnancy (first, second, third, fourth or higher), educational attainment (no education, primary, secondary, post-secondary or higher), occupation type (manual, sales, agricultural, other, not working), wealth index quintiles (specific to urban or rural areas); (2) Women’s agency: whether getting permission and going alone for getting medical help were big problems, person who usually decide the respondent’s healthcare; (3) Healthcare access: whether getting money needed for treatment and the distance to the nearest health facility were big problems, and the type of health facility where the respondent received antenatal care (ANC) (at a public health facility, a private health facility or a non-governmental organization clinic, at home, or no ANC).

### Conceptual framework

Among factors influencing maternal and child health, socio-demographic factors unlikely influence their health outcomes directly. Rather, their influence is most likely mediated through multiple inter-related proximal factors. For example, mothers with higher educational attainment and greater household wealth may be empowered to exercise their agency and actively seek the necessary health services. In this example, adjusting for proximal factors such as women’s agency and healthcare access may reduce the overall effects of the distal factors (socio-demographic).

Following this reasoning, we adapted a conceptual framework recommended by Victora and colleagues [11], based on hypotheses derived from the literature and plausibility reasoning. We hypothesized that TTCV vaccination was influenced by proximal factors related to healthcare access, which were themselves influenced by factors related to women’s agency in seeking care. These, in turn, were influenced by socio-demographic factors. The aim of the conceptual framework is to understand the extent of the role of more distal factors (e.g., socio-demographic variables) on vaccination that is explained by more proximal factors (e.g., healthcare access). Appendix 2 shows a simple schema of the conceptual framework.

A review on maternal immunization in developing countries summarized various socio-economic factors associated with tetanus vaccine uptake by pregnant women in Bangladesh, India, Kenya, and Ethiopia [12]. These factors included mother’s age, birth order, urban or rural residence, educational level, socioeconomic status, woman’s occupation, among the others.

As ANC is the most important strategy to provide maternal immunisation [13], including TTCV, it seems reasonable to assume factors related to healthcare and ANC access as the most proximal to being unvaccinated. Studies have shown that ANC uptake is influenced by many socio-demographic factors. In Ethiopia, an analysis of DHS 2016 data found that higher maternal education status and mothers with greater household wealth were more likely to access ANC while the uptake was low among mothers who had traditional belief and those with five children and more [14]. In addition, a systematic review summarizing the factors influencing ANC uptake showed that most factors were related to socio-demographic domain: maternal education, husband’s education, marital status, household income, mother’s employment, media exposure, having a history of obstetric complications, and parity [15].

Women’s agency is the extent of belief in herself, the awareness of her rights, and make choices to control acquired resources. A 2016 systematic review showed that, in developing countries, women’s empowerment was associated with improved maternal and child health outcomes such as access to ANC and maternal vaccination [16]. A previous analysis of MDHS 2015-16 dataset also demonstrated the association between women empowerment and fewer barriers in accessing healthcare, especially among women with a higher educational attainment, fewer children, from rich households, and those residing in urban areas [17].

### Statistical analysis

We described continuous variables using median and interquartile range; categorical variables using frequency and percentage. Logistic regression models were used in univariable and multivariable analyses accounting for the MDHS sampling weight to investigate factors associated with being unvaccinated. Odds ratios (OR) with their corresponding confidence intervals (CI) were reported. Independent variables were assessed for possible collinearity using Phi and Cramer’s V correlation coefficients (further details can be found in Appendix 4). Multivariable analysis steps and model interpretation under the conceptual framework are summarized in Appendix 3. All analyses were performed using R software (version 4.1.0) [18] and survey analysis was performed using survey package [19]. Correlation coefficients of independent variables were computed using vcd package [20].

### Ethics statement

The 2015-16 MDHS has obtained ethics approval from both ICF Institutional Review Board and Ethics Review Committee of the Department of Medical Research, Ministry of Health and Sports, Myanmar. We obtained the permission from ICF to access the MDHS datasets.

## Results

Our results showed that 69% resided in regions and 77% in rural areas. The median age was 30 years (interquartile range: 26, 36). A third of mothers had their first childbirth in the previous five years. Most mothers (84%) had at least primary school education. Sixteen percent of mothers reported that their decisions of healthcare were made by husband, partner, or others. One in eight mothers did not receive ANC during their last pregnancy (Table 1).

**Table 1.** Characteristics of mothers and the percentage of being unvaccinated with tetanus toxoid containing vaccine.

|  | Number of mothers | % | Unvaccinated* |  |  |
| --- | --- | --- | --- | --- | --- |
|  |  |  | n | % | (95% CI) |
| Overall | 3,582 |  | 990 | 27.6 | (25.1, 30.0) |
| **Socio-demographic** |  |  |  |  |  |
| State or region of residence |  |  |  |  |  |
| Region | 2,452 | 68.5 | 639 | 26.1 | (24.3, 27.9) |
| State | 1,130 | 31.5 | 351 | 31.1 | (28.4, 33.9) |
| Urban or rural residence |  |  |  |  |  |
| Urban | 838 | 23.4 | 163 | 19.5 | (16.9, 22.3) |
| Rural | 2,744 | 76.6 | 827 | 30.1 | (28.4, 31.9) |
| Age of women |  |  |  |  |  |
| 15-24 | 660 | 18.4 | 211 | 32.0 | (28.5, 35.7) |
| 25-29 | 924 | 25.8 | 248 | 26.8 | (24.0, 29.8) |
| 30-34 | 912 | 25.5 | 226 | 24.8 | (22.0, 27.7) |
| 35-49 | 1,087 | 30.3 | 306 | 28.2 | (25.5, 30.9) |
| Birth order of last pregnancy |  |  |  |  |  |
| 1st | 1,222 | 34.1 | 302 | 24.7 | (22.3, 27.3) |
| 2nd | 920 | 25.7 | 220 | 23.9 | (21.2, 26.8) |
| 3rd | 617 | 17.2 | 178 | 28.8 | (25.3, 32.6) |
| 4th or higher | 824 | 23.0 | 290 | 35.2 | (31.9, 38.6) |
| Highest education level of mothers |  |  |  |  |  |
| No education | 587 | 16.4 | 261 | 44.5 | (40.4, 48.6) |
| Primary | 1,629 | 45.5 | 480 | 29.5 | (27.3, 31.8) |
| Secondary | 1,069 | 29.8 | 205 | 19.2 | (16.9, 21.7) |
| More than secondary | 298 | 8.3 | 45 | 15.1 | (11.3, 19.8) |
| Mother's occupation |  |  |  |  |  |
| Manual | 1,060 | 29.6 | 323 | 30.5 | (27.7, 33.4) |
| Sales | 541 | 15.1 | 129 | 23.8 | (20.4, 27.7) |
| Agricultural | 512 | 14.3 | 174 | 34.0 | (29.9, 38.3) |
| Other | 210 | 5.9 | 46 | 21.9 | (16.6, 28.2) |
| Not working | 1,249 | 34.9 | 313 | 25.1 | (22.7, 27.6) |
| (Missing) | 10 | 0.3 |  |  |  |
| Quintiles of woman's wealth index (specific to urban/rural areas) |  |  |  |  |  |
| Lowest | 949 | 26.5 | 341 | 35.9 | (32.9, 39.1) |
| Second | 812 | 22.7 | 229 | 28.2 | (25.2, 31.5) |
| Third | 669 | 18.7 | 171 | 25.6 | (22.3, 29.1) |
| Fourth | 585 | 16.3 | 131 | 22.4 | (19.1, 26.0) |
| Highest | 567 | 15.8 | 119 | 21.0 | (17.8, 24.6) |
| Women's agency |  |  |  |  |  |
| Getting medical help for self: getting permission to go |  |  |  |  |  |
| Not a big problem | 3,407 | 95.1 | 928 | 27.2 | (25.8, 28.8) |
| Big problem | 175 | 4.9 | 62 | 35.4 | (28.5, 43.1) |
| Person who usually decides on woman's healthcare |  |  |  |  |  |
| Self | 1,314 | 36.7 | 355 | 27.0 | (24.6, 29.5) |
| With husband or partner | 1,534 | 42.8 | 424 | 27.6 | (25.4, 30.0) |
| By husband, partner, or others | 575 | 16.1 | 157 | 27.3 | (23.7, 31.2) |
| (Missing) | 159 | 4.4 |  |  |  |
| Getting medical help for self: not wanting to go alone |  |  |  |  |  |
| Not a big problem | 2,408 | 67.2 | 602 | 25.0 | (23.3, 26.8) |
| Big problem | 1,174 | 32.8 | 388 | 33.0 | (30.4, 35.8) |
| Healthcare access |  |  |  |  |  |
| Getting medical help for self: getting money needed for treatment |  |  |  |  |  |
| Not a big problem | 2,192 | 61.2 | 523 | 23.9 | (22.1, 25.7) |
| Big problem | 1,390 | 38.8 | 467 | 33.6 | (31.1, 36.2) |
| Getting medical help for self: distance to health facility |  |  |  |  |  |
| Not a big problem | 2,601 | 72.6 | 624 | 24.0 | (22.4, 25.7) |
| Big problem | 981 | 27.4 | 367 | 37.4 | (34.4, 40.5) |
| Place of antenatal care |  |  |  |  |  |
| Public health facilities | 2,133 | 59.5 | 412 | 19.3 | (17.7, 21.1) |
| Private health facilities or NGO clinics | 289 | 8.1 | 68 | 23.5 | (18.8, 28.9) |
| Home | 565 | 15.8 | 163 | 28.8 | (25.2, 32.8) |
| Mixed | 136 | 3.8 | 32 | 23.5 | (16.9, 31.7) |
| None | 460 | 12.8 | 317 | 68.9 | (64.4, 73.1) |

CI, Confidence Interval
\* Vaccinated mothers include those who has received two injections of tetanus toxoid containing vaccine during the last pregnancy, two or more injections with the last one within three years of the last pregnancy, three or more injections with the last one within five years of the last pregnancy, four or more injections with the last one within 10 years of the last pregnancy, and five or more injections at any time prior to the last pregnancy

### Unvaccinated mothers

Overall, 28% (95% CI: 69%, 74%) of mothers reported that they were unvaccinated. The vast majority of vaccinated mothers received two TTCV injections during the last pregnancy (the first definition in Appendix 1) while fewer than 30 mothers fulfilled the remaining definitions each. At subnational level, the percent unvaccinated was the highest in Shan State (42%), followed by Magway Region (33%), Kayin State (33%), and Sagaing Region (32%) (Figure 1). The percentage of being unvaccinated in each category of explanatory variables is shown in Table 1.

**Figure 1.**
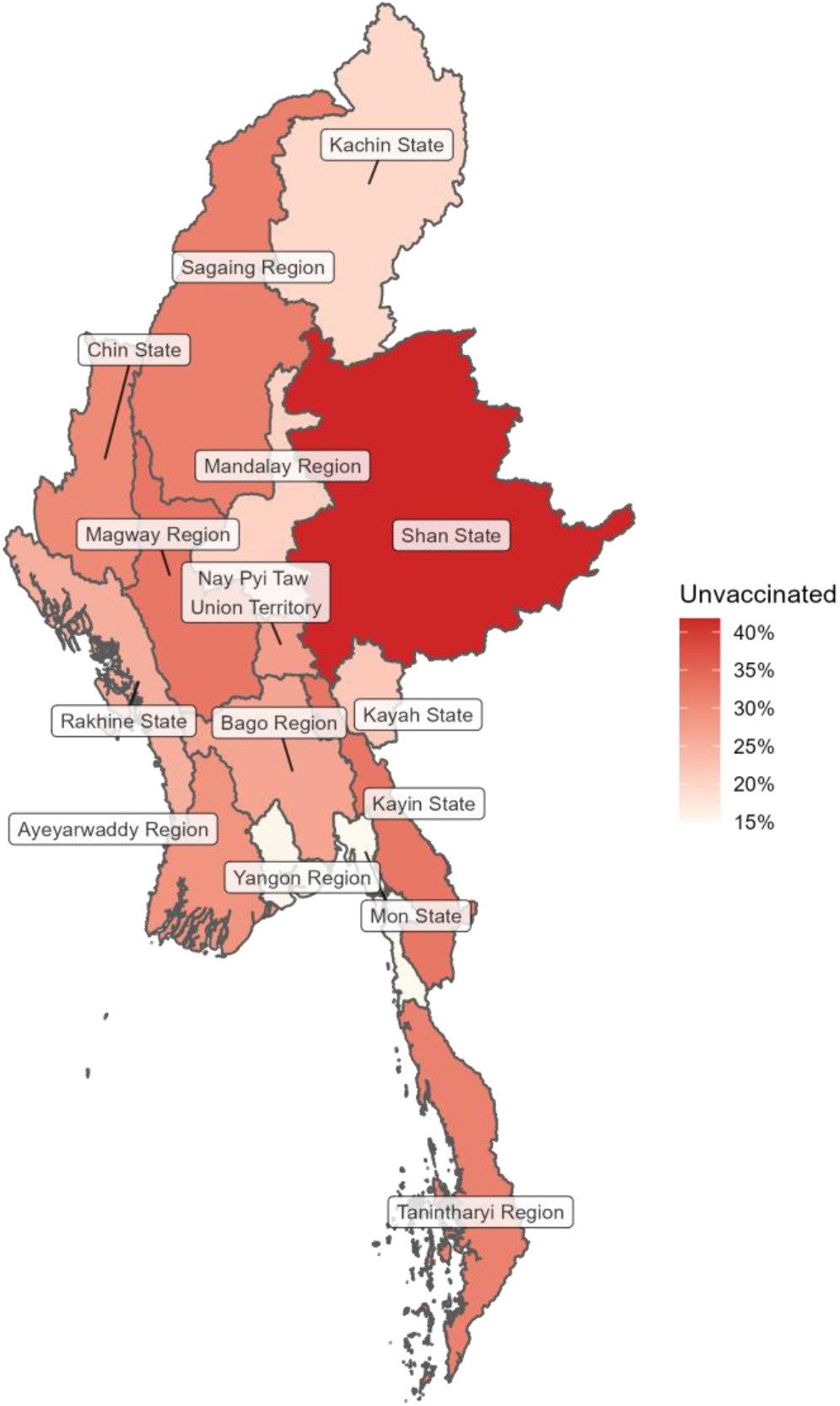
Percentage of unvaccinated mothers with tetanus toxoid containing vaccine in each state or region.

We also estimated that mothers who received at least one dose of TTCV during their last pregnancy was 84% (95%CI: 82%, 87%) with lowest estimates in Shan (69%), Kayin (75%) and Chin (79%) States. Figure 2 shows that the percentage of mothers who missed a second dose of TTCV varied between 5% and 17% at subnational level. Notably, the subnational administrative areas with the highest percentage were regions: Magway (17%), Sagaing (17%), Nay Pyi Taw Union Territory (16%), Bago (16%), and Tanintharyi (14%) (Figure 2).

**Figure 2.**
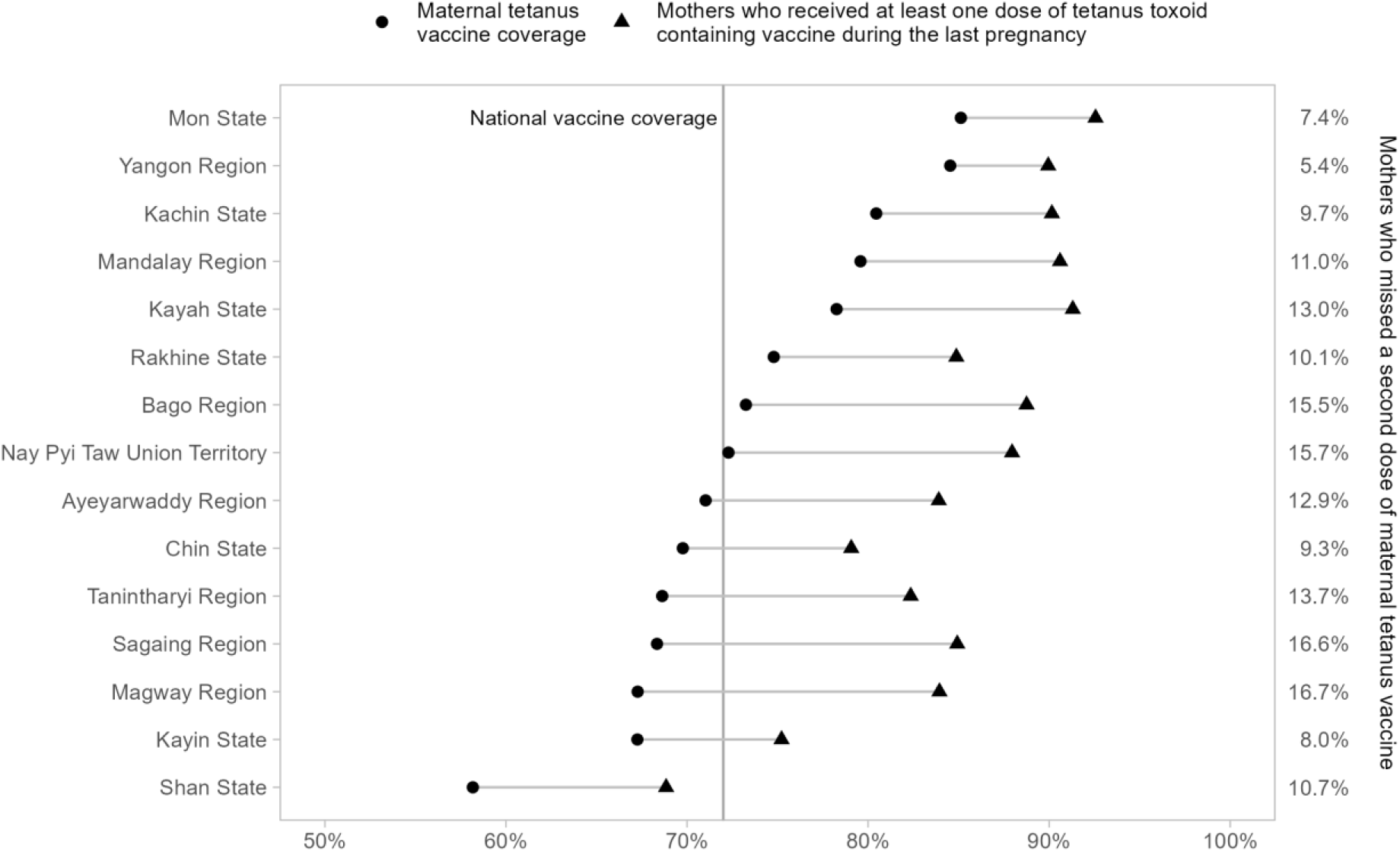
Mothers who missed a second dose of maternal tetanus vaccine in each state and region. Mothers who missed a second dose of maternal tetanus vaccine is the percentage difference between the vaccine coverage and the percentage of mothers who received at least one dose of tetanus toxoid containing vaccine during the last pregnancy

### Factors associated with being unvaccinated

Table 2 shows the results of multivariable analysis under the conceptual framework. Model 1 results represent overall effects of socio-demographic variables. Model 2 provided the effect estimates of women’s agency variables after adjusting for the confounding effects of socio-demographic factors. Model 3 results show the effect estimates of healthcare access factors after adjusting for socio-demographic and women’s agency variables. We found that older age and higher educational attainment of mothers were associated with lower odds of being unvaccinated. On the other hand, not receiving ANC services or receiving ANC in non-public health facilities (private health facilities or non-governmental organization clinic, at home, or mixed, compared to public health facilities) was associated with higher odds of being unvaccinated. Maternal educational attainment remained associated with being unvaccinated after adjusting for the confounding effects of variables in women’s agency and healthcare access domains. Additionally, there was little evidence that maternal age was associated with being unvaccinated in Model 3, suggesting that the effects of maternal age is mediated primarily through mothers’ healthcare access.

**Table 2.**
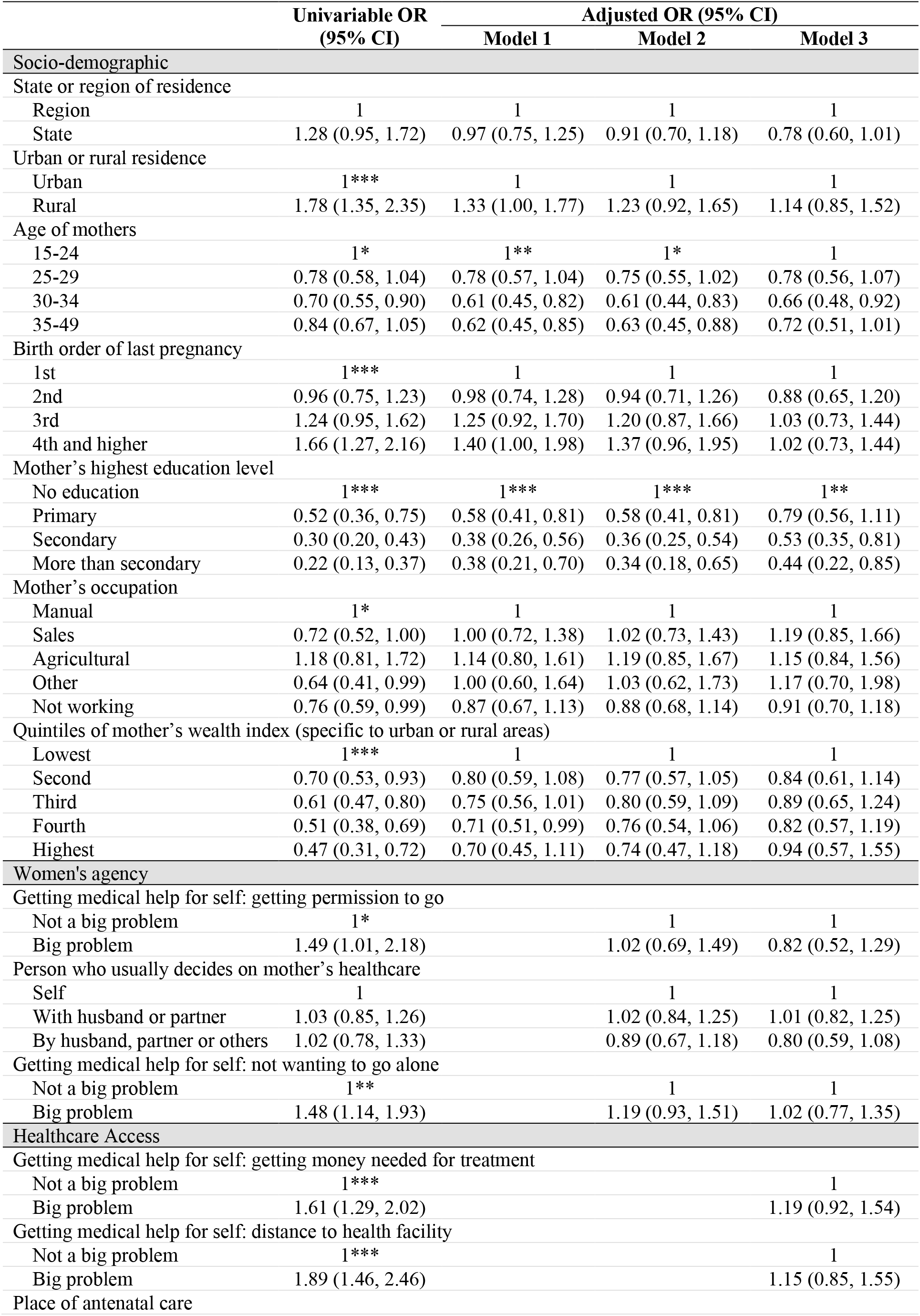
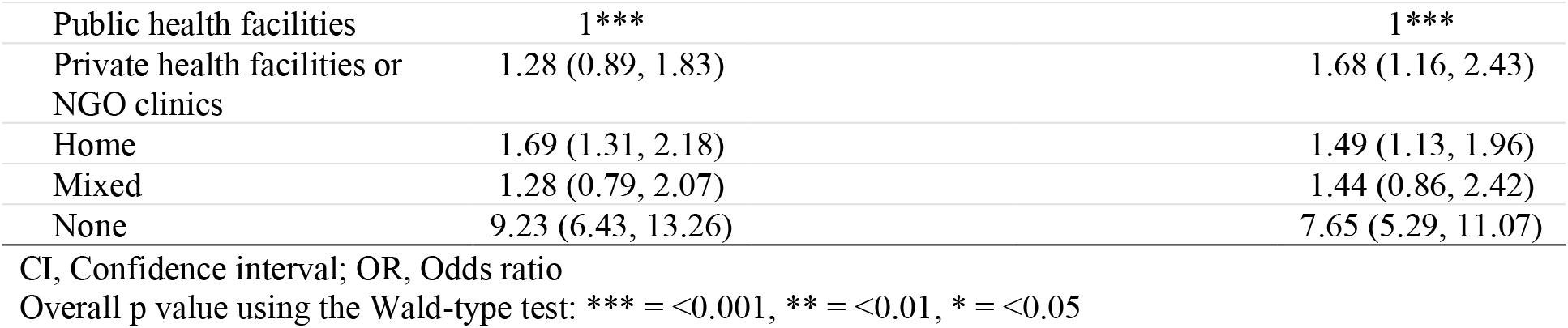
Factors associated with being unvaccinated with tetanus toxoid containing vaccine.

## Discussion

We found important regional variations in the percentage of unvaccinated mothers (ranging from 42% in Shan State to 15% in Mon State), and the percentage of mothers who missed a second dose of TTCV. Notably, we found that the percentage of unvaccinated mothers was higher among younger mothers, and mothers with lower educational attainment. Among these factors, differences in the percentage of unvaccinated mothers maternal age were largely mediated through their access to healthcare and ANC services. Maternal educational attainment remained associated with vaccination status after controlling for healthcare access variables. A lack of ANC or receiving ANC in non-public healthcare facilities were strongly associated with being unvaccinated.

Our analysis showed that the overall percentage of being unvaccinated with TTCV was 28%, translating to the vaccine coverage of 72%. This was lower than the vaccine coverage (88%) officially reported to the WHO by the Myanmar health ministry in 2016 [21]. Although this gap may be explained by methodological differences, it remains a possibility that the official vaccine coverage may underestimate the gap in maternal tetanus vaccination services in the country. In the current analysis, the national vaccine coverage was lower than most countries in the WHO Southeast Asia region in 2016, except Nepal (67%), Indonesia (65%), and Timor-Leste (37%) [21].

We also analysed the percentage of mothers who missed a second dose of TTCV (Figure 2). This information is useful because reasons of not receiving even the first dose of TTCV may differ from reasons of not receiving a second dose. The former is likely due to the lack of access to vaccination services, to information related to these services, or vaccine hesitancy. However, mothers who missed a second dose of TTCV may be due to different reasons, for example, supply side problems: inadequate coverage of vaccination services, poor service quality, frequent stock out, inadequate competent healthcare workers. Our analysis identified regional differences in the percentage of mothers who missed a second dose of TTCV. These percentages are greater in the regions compared to the states. These findings point to an opportunity to significantly improve the vaccine coverage in Myanmar and warrant further investigation.

At subnational level, the percentage of mothers who received at least one dose of TTCV was lowest in Shan, Kayin and Chin States. In addition, the percentage of unvaccinated mothers in these states was among the highest of states and regions (Figure 1). This may be due to limited access to tetanus vaccine. Kayin State and certain areas in Shan State have long been known as chronic conflict-affected areas. Although Myanmar has transitioned to democracy since 2011, conflicts in these areas continued while a multilateral ceasefire agreement involving the military, the newly elected civilian government, and several ethnic armed organizations were being negotiated in parallel [22, 23]. As such, pregnant women residing in these areas faced tremendous challenges in accessing essential maternal health services. Studies aiming to understand the maternal health in these areas are limited. A survey was conducted in 2006 in several accessible conflict-affected areas in Eastern Myanmar (including Shan and Kayin States) near the border with Thailand. The survey found that only 18% and 4% of mothers in Shan and Kayin States, respectively, reported that they have received two doses of tetanus toxoid. The corresponding percentages of mothers attending ANC at least four visits were 45% and 7%, respectively. In addition, women also reported their experience of human rights violations such as destruction or stealing of field and livestock, forced labor, forced relocation, direct physical attacks by soldiers or authorities, and landmines [24]. Separately, Chin State is highly mountainous with few transportation links, which poses challenges for access to maternal health services [25]. A study analyzing MDHS data observed regional differences in ANC coverage (at least four antenatal visits): the coverage in Shan, Kayin and Chin States ranged from 37 to 50%, compared with the coverage in Yangon Region of 82% [26].

Of particular concern is the high percentage of unvaccinated mothers among those who did not access or receive ANC. Approximately 1 in 8 mothers reported not receiving any ANC and of these, more than half were unvaccinated. In Myanmar, ANC services including maternal tetanus vaccination are primarily provided through networks of public health facilities/centres at the village level [10]. Our results show that this system can effectively provide immunization services to most mothers, although the lower coverage among mothers receiving ANC outside the public health sector – private or NGO facilities or at home – points to an opportunity to increase vaccination coverage in these sectors. ANC at home is usually provided by trained midwives and lady health visitors, but in some rural areas ANC is provided by traditional birth attendants who are not trained on vaccination. A study in Krabi Province, Thailand, found that although 91% of traditional birth attendants were aware of injections during ANC at the health center, only half knew that it was for tetanus prevention [27]. A similar knowledge gap may exist among traditional birth attendants in Myanmar. These gaps in ANC provision need to be addressed as Myanmar works towards achieving universal health coverage.

Lower maternal educational attainment was strongly associated with being unvaccinated. The association remained after controlling for healthcare access variables. This suggests a synergistic opportunity for promoting access to both education and healthcare in generating demand for and acceptance of vaccination and improving maternal health outcomes. Lower maternal education is related to lower use of antenatal care services [28], while higher maternal education is also associated with higher uptake of maternal immunisation in several low-income countries, including India [29], Bangladesh [30], Kenya [31], and Malawi [12].

Our analysis has several limitations. In common with other DHS surveys, maternal tetanus vaccination status is self-reported and subject to inaccurate recall. However, a previous study from Brazil showed that self-reported uptake of maternal tetanus vaccination can be higher compared with medical records [32]. In addition, ethnicity and religion could be important determinants of maternal vaccination [33]. However, this information was not available in the survey. Nonetheless, the use of a large, nationally representative, and geographically stratified survey dataset is a major strength.

In our analysis, we were able to identify regional, structural, and individual differences in the percentage of mothers unvaccinated with TTCV. Our findings indicate that further research is necessary to investigate the reasons of mothers not receiving a second dose of TTCV. Myanmar’s progress toward achieving WHO’s recommended target of protecting all pregnant women and babies against tetanus will largely depend on the ability to provide accessible ANC to the substantial fraction of mothers who currently do not currently receive it.

## Data Availability

The 2015-16 DHS Myanmar dataset is accessible upon request from: https://dhsprogram.com/

## Data sharing

The 2015-16 DHS Myanmar dataset is accessible upon request from: https://dhsprogram.com/

## Contributors

ZMT, ZR, and CCT conceived the study. ZR and ZMT obtained the data. ZR and ZMT analysed and interpreted the data. ZMT and ZR wrote the first draft of the report. ZMT, ZR, and CCT contributed to the writing of the report. All authors agreed with report results and conclusions. ZMT and CCT accessed and verified the underlying data.

## Acknowledgements

We thank the Demographic and Health Survey Program for providing data for this analysis.

