## Appendices for "Factors associated with maternal tetanus vaccination in Myanmar: An analysis of Demographic and Health Survey data"

**Appendix 1 – Definitions of a newborn protected against neonatal tetanus based on the Myanmar National Guidelines for Antenatal Care**

A newborn is protected against neonatal tetanus if the mother has received any of the following:

1. Two injections of tetanus toxoid containing vaccine during the last pregnancy
2. Two or more injections, the last one within three years of the last pregnancy
3. Three or more injections, the last one within five years of the last pregnancy
4. Four or more injections, the last one within 10 years of the last pregnancy
5. Five or more injections at any time prior to the last pregnancy

**Appendix 2. Conceptual framework for factors associated with being unvaccinated with tetanus toxoid containing vaccine**

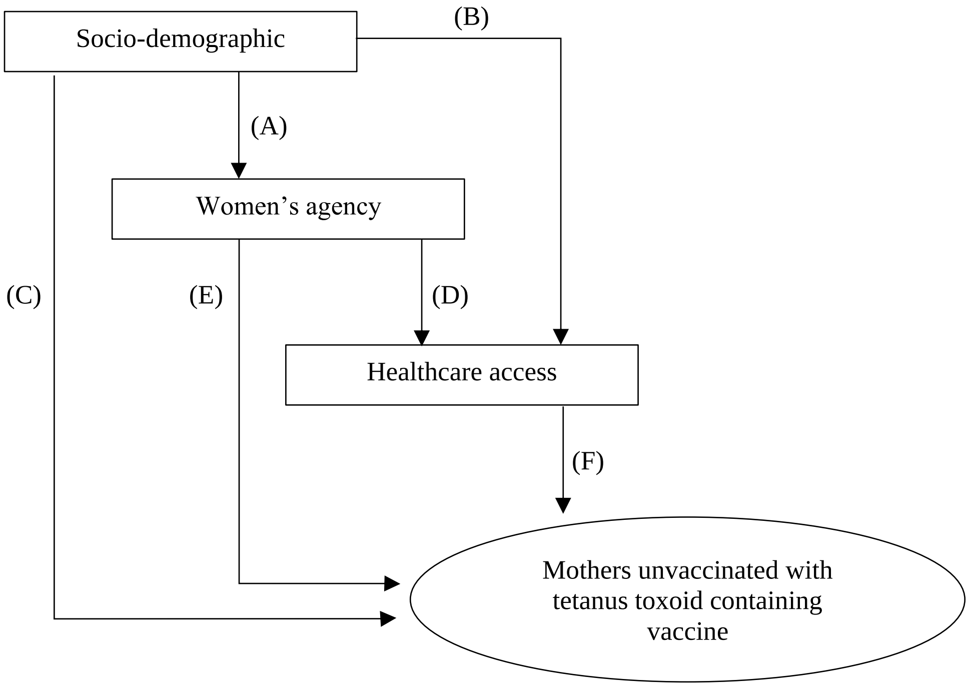

**Appendix 3. Steps of multivariable analysis to identify factors associated with being unvaccinated**

| Model | Explanatory variables | Interpretation | Pathways represented in Figure 1 |
|---|---|---|---|
| 1 | Socio-demographic | Overall effects of socio-demographic variables, without adjusting for the women's agency and healthcare access variables | A + B + C |
| 2 | Socio-demographic + Women's agency | Effects of women's agency variables adjusted for confounding effects of socio-demographic variables | D + E |
| | | Effects of socio-demographic variables that are not mediated through the variables in women's agency domain | B + C |
| 3 | Socio-demographic + Women's agency + Healthcare access | Effects of healthcare access variables adjusted for confounding effects of both socio-demographic and women's agency variables | F |
| | | Effects of women's agency variables that are not mediated through healthcare access variables | E |
| | | Effects of socio-demographic variables that are not mediated through women's agency nor healthcare access variables | C |

**Appendix 4 – Correlation coefficients of independent variables**

Independent variables were assessed for possible collinearity using their correlation coefficients. We used Phi coefficient for two dichotomous variables. This coefficient ranges from -1 to 1 with a value at the extreme ends indicates a greater degree of correlation. If either of the variables consisted of three or more categories, Cramer's V coefficient was used. This coefficient ranges from 0 to 1 – a higher value indicates a greater degree of correlation. Two independent variables with a Phi coefficient of either <-0.8 or >0.8 and a Cramer's V coefficient of >0.8 was considered highly correlated, indicating a possible collinearity. The table below shows that the independent variables are not highly correlated.

| First variable | Second variable | Correlation measure | Coefficient |
|---|---|---|---|
| State or region of residence | Urban or rural residence | Phi | 0.06 |
| State or region of residence | Age of women | Cramer's V | 0.063 |
| State or region of residence | Birth order of last pregnancy | Cramer's V | 0.127 |
| State or region of residence | Highest education level of women | Cramer's V | 0.213 |
| State or region of residence | Women's occupation | Cramer's V | 0.132 |
| State or region of residence | Quintiles of woman's household wealth index | Cramer's V | 0.069 |
| State or region of residence | Getting medical help for self: getting permission to go | Phi | 0.114 |
| State or region of residence | Person who usually decides on woman's healthcare | Cramer's V | 0.068 |
| State or region of residence | Getting medical help for self: not wanting to go alone | Phi | 0.13 |
| State or region of residence | Getting medical help for self: getting money needed for treatment | Phi | 0.141 |
| State or region of residence | Getting medical help for self: distance to health facility | Phi | 0.136 |
| State or region of residence | Place of antenatal care | Cramer's V | 0.117 |
| Urban or rural residence | Age of women | Cramer's V | 0.036 |
| Urban or rural residence | Birth order of last pregnancy | Cramer's V | 0.17 |
| Urban or rural residence | Highest education level of women | Cramer's V | 0.384 |
| Urban or rural residence | Women's occupation | Cramer's V | 0.302 |
| Urban or rural residence | Quintiles of woman's household wealth index | Cramer's V | 0.016 |
| Urban or rural residence | Getting medical help for self: getting permission to go | Phi | 0.09 |
| Urban or rural residence | Person who usually decides on woman's healthcare | Cramer's V | 0.069 |
| Urban or rural residence | Getting medical help for self: not wanting to go alone | Phi | 0.147 |
| Urban or rural residence | Getting medical help for self: getting money needed for treatment | Phi | 0.138 |
| Urban or rural residence | Getting medical help for self: distance to health facility | Phi | 0.214 |
| Urban or rural residence | Place of antenatal care | Cramer's V | 0.284 |
| Age of women | Birth order of last pregnancy | Cramer's V | 0.354 |

| | | | |
|---|---|---|---|
| Age of women | Highest education level of women | Cramer's V | 0.126 |
| Age of women | Women's occupation | Cramer's V | 0.068 |
| Age of women | Quintiles of woman's household wealth index | Cramer's V | 0.037 |
| Age of women | Getting medical help for self: getting permission to go | Cramer's V | 0.03 |
| Age of women | Person who usually decides on woman's healthcare | Cramer's V | 0.042 |
| Age of women | Getting medical help for self: not wanting to go alone | Cramer's V | 0.045 |
| Age of women | Getting medical help for self: getting money needed for treatment | Cramer's V | 0.036 |
| Age of women | Getting medical help for self: distance to health facility | Cramer's V | 0.048 |
| Age of women | Place of antenatal care | Cramer's V | 0.045 |
| Birth order of last pregnancy | Highest education level of women | Cramer's V | 0.224 |
| Birth order of last pregnancy | Women's occupation | Cramer's V | 0.093 |
| Birth order of last pregnancy | Quintiles of woman's household wealth index | Cramer's V | 0.123 |
| Birth order of last pregnancy | Getting medical help for self: getting permission to go | Cramer's V | 0.106 |
| Birth order of last pregnancy | Person who usually decides on woman's healthcare | Cramer's V | 0.027 |
| Birth order of last pregnancy | Getting medical help for self: not wanting to go alone | Cramer's V | 0.077 |
| Birth order of last pregnancy | Getting medical help for self: getting money needed for treatment | Cramer's V | 0.213 |
| Birth order of last pregnancy | Getting medical help for self: distance to health facility | Cramer's V | 0.155 |
| Birth order of last pregnancy | Place of antenatal care | Cramer's V | 0.149 |
| Highest education level of women | Women's occupation | Cramer's V | 0.275 |
| Highest education level of women | Quintiles of woman's household wealth index | Cramer's V | 0.254 |
| Highest education level of women | Getting medical help for self: getting permission to go | Cramer's V | 0.118 |
| Highest education level of women | Person who usually decides on woman's healthcare | Cramer's V | 0.071 |
| Highest education level of women | Getting medical help for self: not wanting to go alone | Cramer's V | 0.186 |
| Highest education level of women | Getting medical help for self: getting money needed for treatment | Cramer's V | 0.269 |
| Highest education level of women | Getting medical help for self: distance to health facility | Cramer's V | 0.23 |
| Highest education level of women | Place of antenatal care | Cramer's V | 0.236 |
| Women's occupation | Quintiles of woman's household wealth index | Cramer's V | 0.127 |
| Women's occupation | Getting medical help for self: getting permission to go | Cramer's V | 0.17 |
| Women's occupation | Person who usually decides on woman's healthcare | Cramer's V | 0.08 |
| Women's occupation | Getting medical help for self: not wanting to go alone | Cramer's V | 0.177 |
| Women's occupation | Getting medical help for self: getting money needed for treatment | Cramer's V | 0.202 |
| Women's occupation | Getting medical help for self: distance to health facility | Cramer's V | 0.207 |

| | | | |
|---|---|---|---|
| Women's occupation | Place of antenatal care | Cramer's V | 0.132 |
| Quintiles of woman's household wealth index | Getting medical help for self: getting permission to go | Cramer's V | 0.102 |
| Quintiles of woman's household wealth index | Person who usually decides on woman's healthcare | Cramer's V | 0.064 |
| Quintiles of woman's household wealth index | Getting medical help for self: not wanting to go alone | Cramer's V | 0.162 |
| Quintiles of woman's household wealth index | Getting medical help for self: getting money needed for treatment | Cramer's V | 0.336 |
| Quintiles of woman's household wealth index | Getting medical help for self: distance to health facility | Cramer's V | 0.219 |
| Quintiles of woman's household wealth index | Place of antenatal care | Cramer's V | 0.143 |
| Getting medical help for self: getting permission to go | Person who usually decides on woman's healthcare | Cramer's V | 0.1 |
| Getting medical help for self: getting permission to go | Getting medical help for self: not wanting to go alone | Phi | 0.25 |
| Getting medical help for self: getting permission to go | Getting medical help for self: getting money needed for treatment | Phi | 0.233 |
| Getting medical help for self: getting permission to go | Getting medical help for self: distance to health facility | Phi | 0.279 |
| Getting medical help for self: getting permission to go | Place of antenatal care | Cramer's V | 0.124 |
| Person who usually decides on woman's healthcare | Getting medical help for self: not wanting to go alone | Cramer's V | 0.062 |
| Person who usually decides on woman's healthcare | Getting medical help for self: getting money needed for treatment | Cramer's V | 0.042 |
| Person who usually decides on woman's healthcare | Getting medical help for self: distance to health facility | Cramer's V | 0.062 |
| Person who usually decides on woman's healthcare | Place of antenatal care | Cramer's V | 0.073 |
| Getting medical help for self: not wanting to go alone | Getting medical help for self: getting money needed for treatment | Phi | 0.397 |
| Getting medical help for self: not wanting to go alone | Getting medical help for self: distance to health facility | Phi | 0.585 |
| Getting medical help for self: not wanting to go alone | Place of antenatal care | Cramer's V | 0.17 |
| Getting medical help for self: getting money needed for treatment | Getting medical help for self: distance to health facility | Phi | 0.48 |
| Getting medical help for self: getting money needed for treatment | Place of antenatal care | Cramer's V | 0.176 |
| Getting medical help for self: distance to health facility | Place of antenatal care | Cramer's V | 0.247 |

1
